# Potential Causal Relationship between Faster Walking Pace and Reduced Migraine Risk: A Mendelian Randomization Study

**DOI:** 10.1101/2024.08.07.24311600

**Authors:** Xueen Liu, Jiale Zhang

## Abstract

**Objectives:** Prior observational studies have suggested a potential association between the usual walking pace and migraine. In the present study, we utilized Mendelian randomization (MR) to investigate the presence of causality and elucidate the specific causal relationship between these two variables.

**Methods:** We performed a genome-wide association study on a population of 499,562 individuals of European ancestry, which revealed 34 genetic variants that exhibited a strong association with the usual walking pace. Additionally, we obtained summary statistics for genome-wide association studies on migraine from several sources. To assess the causal estimates, we employed the random effects inverse variance weighted method (IVW) and several other Mendelian randomizations (MR) methods, including MR-Egger, weighted median, Simple mode, Weighted mode, and MR-PRESSO, to confirm the robustness of our results.

**Results:** Our analysis demonstrated a strong causal association between genetically predicted usual walking pace and a decreased risk of migraine, as determined by inverse variance weighted analysis (odds ratio = 0.33; 95% CI = 0.17 to 0.63; *P* < 0.001). This association was consistently observed across our investigation’s various Mendelian randomization (MR) methods.

**Conclusions:** This study supports a potential causal association between increased walking speed and a decreased risk of migraine.

**Highlight:**

1. This study used MR to investigate the causal relationship between the usual walking pace and migraine.
2. A genome-wide association study identified 34 genetic variants strongly associated with the usual walking pace in a European population.
3. Multiple MR methods confirmed a consistent causal association between genetically predicted usual walking pace and decreased migraine risk.
4. Increasing walking speed may reduce the risk of migraine.

## Introduction

Mendelian randomization (MR) is an epidemiological technique that employs genetic variation as an instrumental variable to address confounding factors and reverse causation[1]. By randomly assigning genotypes at conception, MR eliminates the possibility of confounding and reverse causality by Mendel’s second law[2]. As such, MR is widely regarded as a highly reliable tool for predicting causality, as demonstrated in Figure 1. The present study used MR analysis to explore the potential relationship between walking pace and migraine, with genetically predicted walking pace as the primary exposure variable.

**Figure 1.**
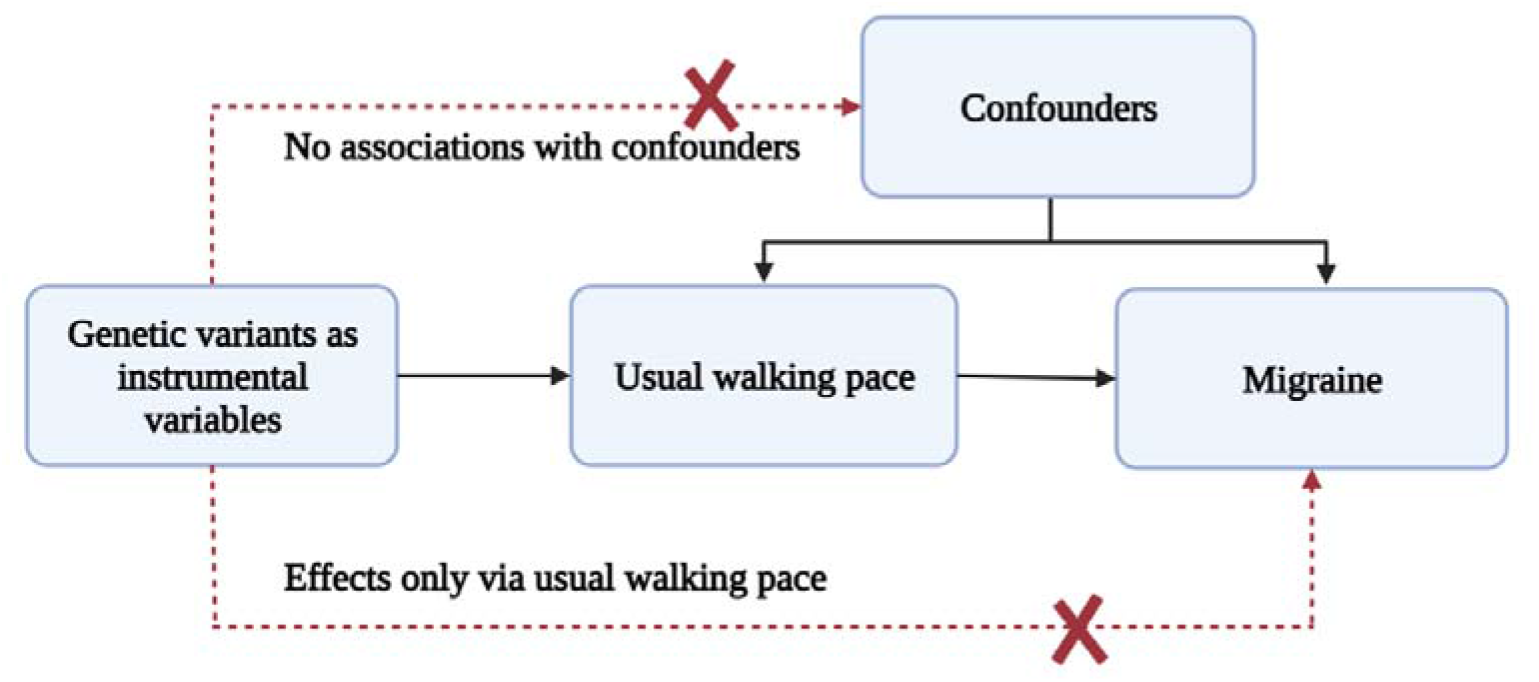
Mendelian randomization analyses to investigate associations between the usual walking pace and the risk of the usual walking pace. the broken lines signify potential causal effects (pleiotropic or direct) between variables that would violate the assumptions of Mendelian randomization.

## Materials and methods

### Study design

To investigate the causal association between the risk of migraine and the usual walking pace, our study employed a two-sample Mendelian randomization approach. The study adhered to the guidelines for the Preferred Reporting of Observational Studies in Epidemiology Using Mendelian Randomization (The STROBE-MR Statement)[3]. Our investigations utilized publicly available genome-wide association study (GWAS) summary data, which had undergone ethical review board approvals prior to their inclusion in our study.

### Instruments selection for usual walking pace

We obtained genome-wide association study (GWAS) summary statistics on the usual walking pace from the UK Biobank, which included 499,562 participants of European ancestry. This dataset was the largest GWAS on the usual walking pace available during our Mendelian randomization (MR) analysis. In the original GWAS study, the usual walking pace was assessed using a touchscreen question that asked participants to describe their usual walking pace as “slow”, “steady/average”, or “brisk”. Further information was provided to participants, which defined a slow pace as less than 3 miles per hour, a steady/average pace as between 3-4 miles per hour, and a brisk pace as more than 4 miles per hour.

As genetic instruments, we selected 46 single nucleotide polymorphisms (SNPs) strongly associated with the usual walking pace at a genome-wide significance level (p < 5×10−8. To maintain independence, we performed a linkage disequilibrium (LD) test (r2 < 0.001 and distance > 10 000 kb) [4]. We did not use palindromic SNPs (rs11125819, rs12127073, rs2054080, rs2132828, rs45583845, rs55929017, rs72636674, rs8005131) with intermediate allele frequencies that pair with each other since they may invert the direction of a causal effect. We utilized proxy SNPs (rs1269454, rs7923609, rs10846920, rs79095373) as substitutes for SNPs not included in the outcome data by searching SNIPA (https://snipa.helmholtz-muenchen.de/snipa3/index.php). We performed a manual inspection using PhenoScanner for screening association with confounder[5]. In the initial analysis, we employed 36 independent SNPs as instrumental variables. The R^2^ was then utilized to evaluate the extent to which each SNP accounted for the variance in the usual walking pace[6]. To evaluate the effectiveness of instrumental variables, we utilized the F statistic, considering an F value greater than 10 as indicative of strong instruments[7].

### Genetic Associations with Migraine

Our GWAS summary statistics for migraine were obtained from the FinnGen project, which could be accessed at https://gwas.mrcieu.ac.uk/datasets/finn-b-G6_MIGRAINE/. There were 8,547 cases of Migraine and 176,107 people who served as controls in this study.

### Statistical analysis

We conducted Mendelian randomization (MR) analyses using R software version 4.2.2 and leveraged the Mendelian Randomization, RadialMR, and MR-PRESSO packages for the MR analyses. The primary MR analyses were conducted using inverse variance weighted (IVW) analyses[8]. The Cochran Q test was employed with a significance level of P < 0.05 to assess the heterogeneity of IVW estimates[9]. Additionally, several sensitivity analyses were performed to ensure the robustness of the results. Specifically, we employed five widely-used MR methods: MR-Egger, weighted median, Simple mode, Weighted mode, and MR-PRESSO. The MR-Egger method, a prominent approach for detecting and correcting pleiotropy, was utilized to estimate the potential impact of pleiotropy on the results. An MR-Egger intercept with a P-value of less than 0.05 indicates the presence of directional pleiotropy, which may result in biased MR estimates[10]. The weighted median approach was utilized to generate a more reliable estimate even when up to 50% of the instruments were invalid, by taking the median of all possible pairwise IVW estimates. Furthermore, two additional MR methods, Simple and Weighted modes, were employed to strengthen the findings’ validity further. The Simple mode selects SNPs based on their association with the exposure and outcome. In contrast, the Weighted mode selects SNPs based on their overall strength of association with the exposure and outcome[8]. These methods provide alternative instrumental variable selection criteria and estimation techniques. Additionally, the MR-PRESSO method, which detects and corrects for outliers and horizontal pleiotropy, was employed to assess the potential impact of outliers on the results[11]. We also use RadialMR to identify outliers. RadialMR is a Mendelian randomization method that improves the accuracy of causal effect estimates by projecting weak instrumental variables onto a vector that maximizes the variance of the exposure variable[12]. It can be applied to high-dimensional genetic data and is useful for large-scale MR studies. For each MR method, we estimated the odds ratios (OR) per one standard deviation (SD) increase in genetically predicted continuous traits, along with corresponding 95% confidence intervals (CI), to determine the causal effect of usual walking pace. A significance threshold of *P* < 0.05 was set to determine statistical significance. Furthermore, we utilized leave-one-out analyses to evaluate the influence of individual SNPs on the overall causal estimates. This approach involves systematically eliminating one SNP at a time and re-analyzing the data to assess the sensitivity of the results to each SNP.

## Results

Our Mendelian randomization analysis utilizing the inverse variance weighted (IVW) method revealed a robust causal association between genetically predicted faster usual walking pace and a decreased risk of migraine (odds ratio = 0.33; 95% CI = 0.17 to 0.663; *P* < 0.001). This effect was consistently observed across most Mendelian randomization methods, as illustrated in Figure 2. Notably, 2 outliers (rs784257, rs7896518) were identified during RadialMR analysis and subsequently excluded from our investigation. 34 SNPs were utilized as genetic instruments, all with an F statistic greater than 39, indicating a strong association with usual walking pace. Our results were unlikely confounded by heterogeneity or pleiotropic effects, as demonstrated by the absence of significant heterogeneity of IVW estimates (Q-test, *P* > 0.98) and directional pleiotropy (MR-Egger intercept, *P* > 0.25). Based on our leave-one-out analyses (Figure 3), we found no significant differences in the estimated causal effects when individual SNPs were excluded, underscoring the robustness of our Mendelian randomization analysis. Consequently, the estimated effect cannot be attributed to any specific SNP. Our study provides evidence suggesting that the genetically predicted usual walking pace may confer a protective effect against the risk of migraine.

**Figure 2.**
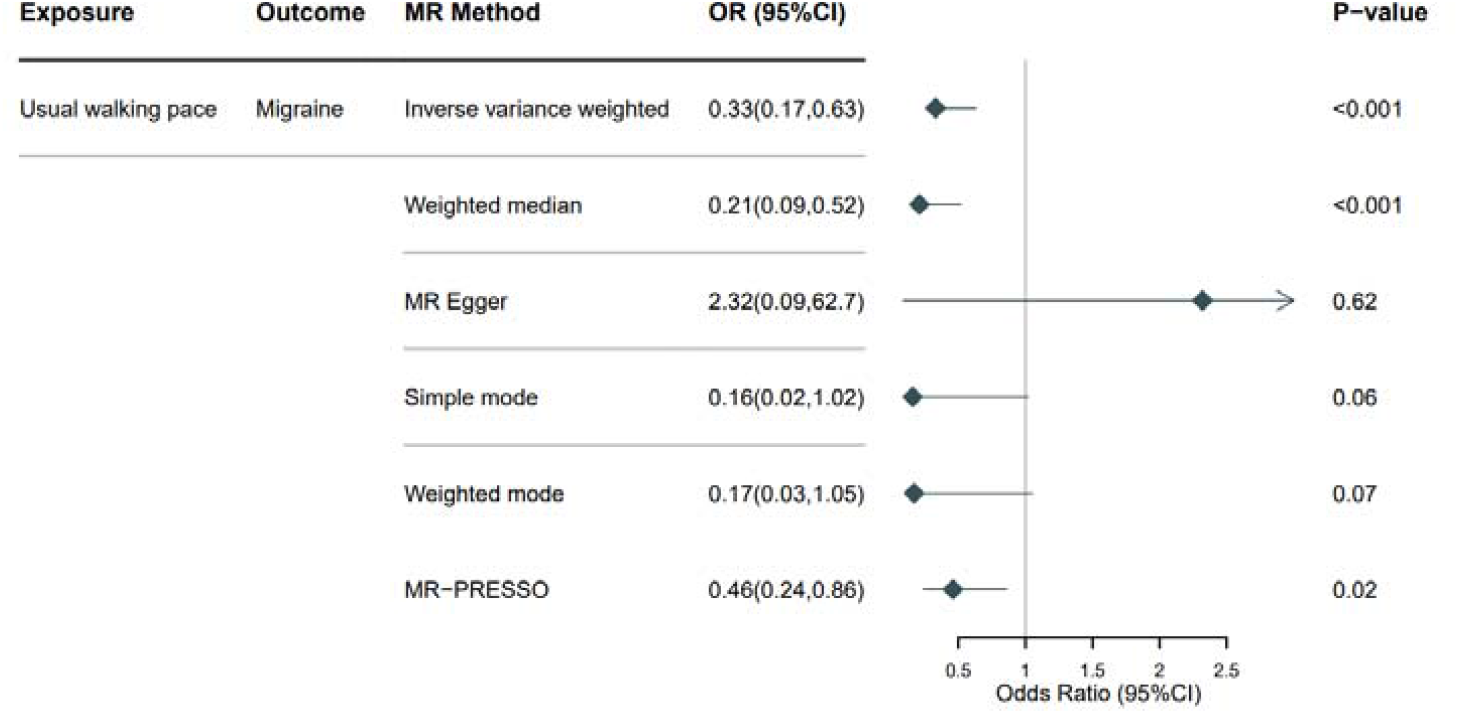
The odds ratios (OR) depicting the associations between genetically predicted usual walking pace and Migraine risk are presented. CI = confidence interval; MR = Mendelian randomization.

**Figure 3.**
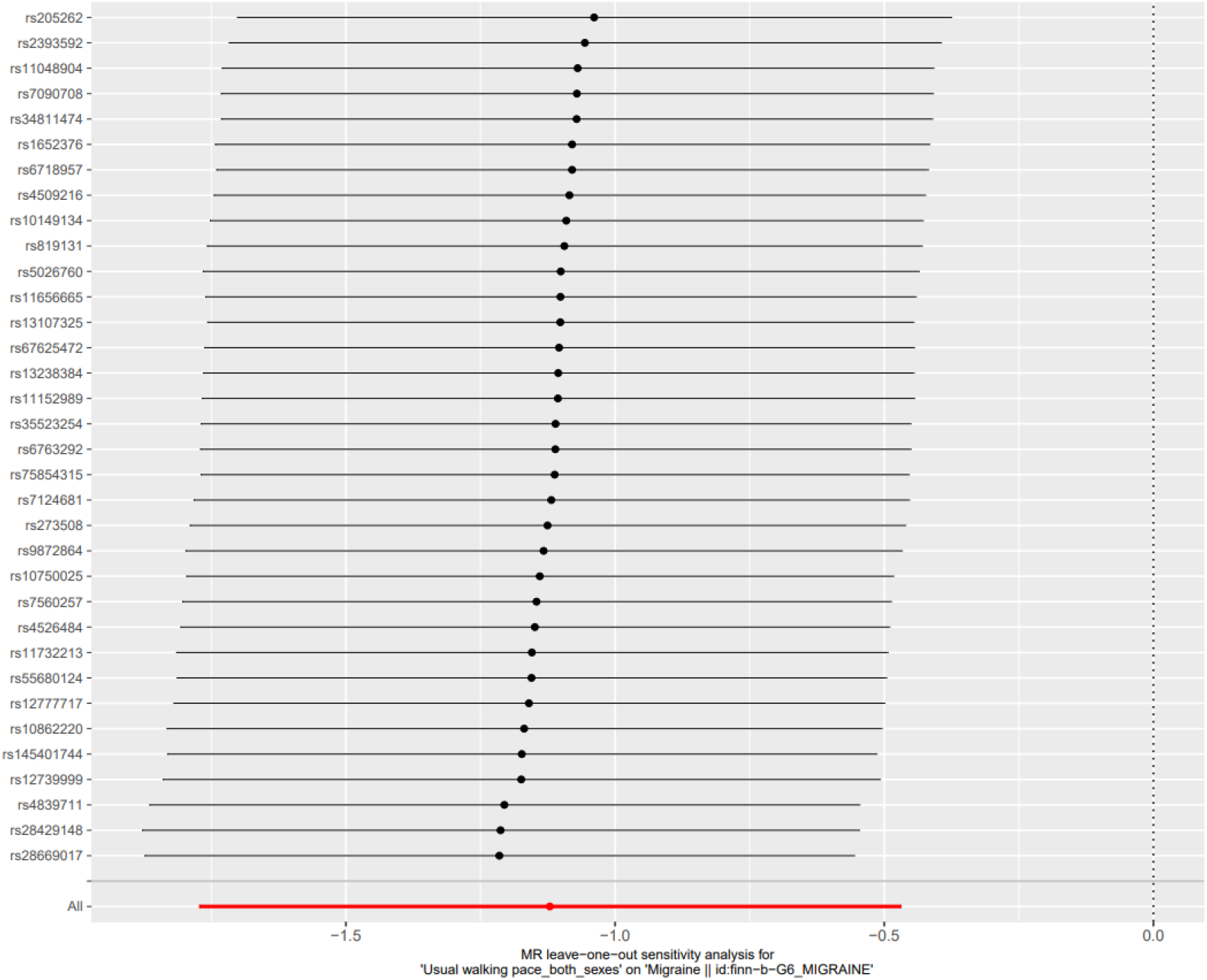
The leave-one-out analyses revealed no significant differences in the estimated causal effects when individual SNPs were removed from our MR analysis.

## Discussion

This study employed a two-sample Mendelian randomization (MR) approach to investigate the causal relationship between habitual walking speed and migraine risk. We followed the guidelines outlined in the STROBE-MR statement and conducted our analysis using publicly available summary statistics from genome-wide association studies (GWAS). Our study population comprised 499,562 participants of European ancestry, making it the largest available GWAS study on walking speed. We utilized multiple MR methods and obtained consistent results, indicating a significant negative correlation between genetically predicted habitual walking speed and migraine risk (OR=0.33, 95% CI=0.17-0.63, P<0.001). Our Mendelian randomization analysis provides compelling evidence that faster habitual walking speed may be a promising modifiable factor for preventing migraine. Additionally, our study results provide further evidence for the role of aerobic exercise in the prevention and management of migraine, reinforcing previous observational studies and emphasizing the importance of incorporating exercise adjustments in migraine prevention and management. However, the exact protective mechanism of habitual walking pace in migraine remains to be fully elucidated.

Our study also has its limitations. Firstly, a notable drawback of MR is it’s potential for horizontal pleiotropy, which may lead to biased causal estimates. However, the consistency of our results with those obtained from alternative MR methods less susceptible to the influence of horizontal pleiotropy provides evidence for our findings. Secondly, we could not perform subgroup analyses due to the need for detailed clinical subtyping information of migraines. Lastly, our study was limited to individuals of European ancestry, and it should be noted that our findings may need to be more applicable to populations of different ancestries. As our study results only report a causal relationship between increased walking pace and decreased migraine risk, further research is needed to address potential mechanisms.

## Author Contribution

JL Z conceived the research idea. XE L conducted the literature search, while XE L wrote the manuscript. JL Z performed software operations and created the figures, and XE L contributed to the final version. All authors reviewed and edited the manuscript and contributed to the analysis through constructive discussions.

## Data Availability Statement

The summary statistics for relative carbohydrate intake and AF are available at the Social Science Genetic Association Consortium (SSGAC; https://www.thessgac.org/data) and IEU open gwas project, respectively.

## Ethics declarations

### Ethics approval and consent to participate

Not applicable.

### Consent for publication

Not applicable.

